# Validation and implementation of a direct RT-qPCR method for rapid screening of SARS-CoV-2 infection by using non-invasive saliva samples

**DOI:** 10.1101/2020.11.19.20234245

**Authors:** Pedro Brotons, Amaresh Perez-Argüello, Cristian Launes, Francesc Torrents, Jesica Saucedo, Joana Claverol, Juan Jose Garcia-Garcia, Gil Rodas, Vicky Fumado, Iolanda Jordan, Eduard Gratacos, Quique Bassat, Carmen Muñoz-Almagro

## Abstract

**Background:** There is an urgent need to curb COVID-19 pandemic through early identification of asymptomatic but infectious cases. We aimed to validate and implement an optimised screening method for detection of SARS-CoV-2 RNA combining use of self-collected raw saliva samples, single-step heat-treated virus inactivation and RNA extraction, and direct RT-qPCR.

**Methods and findings:** The study was conducted in Sant Joan de Deu University Hospital (Barcelona, Spain), including: i) analytical validation against standard RT-qPCR in saliva samples; ii) diagnostic validation against standard RT-qPCR using paired saliva-nasopharyngeal samples obtained from asymptomatic teenagers and young and older adults in a youth sports academy; and iii) high throughput pilot screening of asymptomatic health workers and other staff in the study site.

The proposed method had comparable analytical performance to standard RT-qPCR in saliva. Diagnostic validation included saliva samples self-collected with supervision by 173 participants during 9-12 weeks and nasopharyngeal samples collected from them. At baseline, all participants (100.0%) were negative for SARS-CoV-2 in both paired saliva-nasopharyngeal samples. In the following weeks, standard RT-qPCR yielded 23 positive results in nasopharyngeal samples whereas paired saliva specimens yielded 22 (95.7%) positive and one inconclusive result.

A total of 2,709 participants engaged in the pilot screening, with high rate of participation (83.4% among health workers). Only 17 (0.6%) of saliva samples self-collected by participants in an unsupervised manner were invalid. Saliva was positive in 24 (0.9%) out of 2,692 valid specimens and inconclusive in 27 (1.0%). All 24 saliva-positive participants and 4 with saliva inconclusive results were positive by standard RT-qPCR in nasopharyngeal samples. The pilot showed potential for rapid analytical workflow (up to 384 batched samples can be processed in <2 hours).

**Conclusion:** Direct RT-qPCR on self-collected raw saliva is a simple, rapid, and accurate method with potential to be scaled up for enhanced SARS-CoV-2 community-wide screening.

## Introduction

The burden and health, educational, and economic implications of the Coronavirus Infectious Disease 19 (COVID-19) pandemic have underlined an urgent need for rapid and accurate diagnostics for severe acute respiratory syndrome coronavirus 2 (SARS-CoV-2) [1,2]. Early identification of SARS-CoV-2 is challenging since a proportion of infected individuals can show little or no symptoms for an indeterminate period of time [3,4]. Noticeable rates of asymptomatic and pre-symptomatic infection have been observed across different studies, ranging from 3 to 67% [5]. Asymptomatic or pre-symptomatic individuals are nevertheless likely to be infectious [6,7].

Upper respiratory tract (URT) samples are the specimens currently recommended for diagnosis of COVID-19 [8]. Reverse transcription real-time polymerase chain reaction (RT-qPCR) constitutes the preferred method for detection of SARS-CoV-2, given its high sensitivity and specificity [9]. RT-qPCR accuracy may vary depending on URT sample quality and time elapsed since virus acquisition [10]. Standard RT-qPCR protocols for SARS-CoV-2 typically follow three sequential phases: i) URT sample swabing and sample transport in viral inactivation transport medium (VITM) to the laboratory for analysis or, alternatively, sample transport in viral transport medium (VTM) and inactivation in the laboratory; ii) RNA extraction, purification, and concentration with use of targeted reagents and automated robots; and iii) viral RNA amplification and detection in thermal cyclers. RNA extraction, purification and concentration are slow and cumbersome activities that take from 40 minutes to 3 hours, depending on the type of RNA extraction robot utilised and the number of samples batched together. During the first wave of the pandemic, shortage of personal protection equipment (PPE), swabs, VITM, and RT-PCR reagent supplies created serious bottlenecks in the diagnostic workflow of clinical and epidemiological surveillance laboratories [11].

Saliva appears to be a promising URT specimen type for screening, diagnosis, follow up, and infection control of SARS-CoV-2 in all age groups. Diverse studies have reported consistent detection of SARS-CoV-2 RNA in saliva of symptomatic COVID-19 patients and sensitivities of saliva-based RT-qPCR ranging from 84 to100% compared to paired positive nasopharyngeal (NP) samples [12-14]. While collection of NP or oropharyngeal samples is inconvenient for patients and exposes health care workers to infection risk, saliva specimens can be repeatedly collected or self-collected in a simple, safe, and inexpensive manner without specific training or use of PPEs. In addition, good saliva stability at room temperature can simplify sample transport, avoiding cold-chain conditions [15]. Recently, the U.S. Food and Drug Administration granted accelerated emergency use authorization for the use of saliva, in addition to other respiratory specimen types, to facilitate mass screening of SARS-CoV-2 [16]. However, there is scarce evidence on the implementation of saliva-based screening approaches to identify asymptomatic subjects.

We have developed a novel screening method for SARS-CoV-2 that combines use of self-collected raw saliva samples, heat-treated virus inactivation and RNA extraction in a single step, and RT-qPCR, herein referred as direct RT-qPCR. This simple, safe, and rapid method circumvents use of collection swabs, VITM, and RNA extraction reagents, as well as RNA purification and concentration steps, allows utilisation of different commercial RT-qPCR kits, and minimises dependence on the supply chain of reagents and consumables. The objective of the study was to validate and implement direct RT-qPCR on self-collected saliva for first-line screening of SARS-CoV-2 infection.

## Methods

### Study design and participants

The study was conducted in the Molecular Microbiology Department of Sant Joan de Déu Hospital (SJDH), a university reference maternal and child health medical centre located in Barcelona (Spain), in several successive phases:

#### Phase 1. Analytical validation

SARS-CoV-2 RNA detection yield was assessed in saliva samples by direct RT-qPCR and a 3-phased standard RT-qPCR protocol. Additionally, intra- and inter-assay precision and effect of saliva storage under different conditions on performance were evaluated.

#### Phase 2. Diagnostic validation

Performance of direct RT-qPCR was compared against standard RT-pPCR on paired saliva-NP samples serially obtained from asymptomatic young and adult participants included in a prospective cohort. Outcomes sought were diagnostic sensitivity, specificity, and predictive values of saliva-based direct RT-qPCR against standard RT-qPCR in NP samples.

#### Phase 3. Pilot screening programme

Once validated, saliva-based direct RT-qPCR was deployed in SJDH to screen volunteer health workers and other staff. Planned outcomes were rate of participation (as a proxy for pilot acceptance), identification of positive cases for prevention of COVID-19 nosocomial outbreaks in the setting, and feasibility of unsupervised saliva self-collection by end-users.

### Analytical validation procedures

#### Comparative performance of direct RT-qPCR and standard RT-qPCR in saliva

Samples required for analytical validation were voluntarily provided by healthy adult researchers involved in the study or obtained from SJDH’s Biobank, a research biorepository integrated into the Spanish Biobank Network of Instituto de Salud Carlos III. SARS-CoV2 RNA positive saliva samples or volumes of 90 µL of saliva spiked with 10 µL of NP positive samples were processed by direct RT-qPCR and standard RT-qPCR. Direct RT-PCR workflow involved saliva incubation in block heater for 15 minutes at 96°C to maximise virus inactivation and RNA extraction. RNA amplification was performed using two RT-qPCR kits (GeneFinder^®^ COVID-19 Plus RealAmp kit, Elitech, France; TaqPath^®^ COVID-19 RT-PCR kit, Thermofisher, US) and two thermal cycler platforms (Applied Biosystems^®^ QuantStudio 7 and Applied Biosystems^®^ Prism 7500, Thermofisher, US). Standard RT-qPCR workflow included viral chemical inactivation and RNA extraction, purification, and concentration using NucliSense^®^ easyMAG^®^ platform and reagents (bioMérieux, The Netherlands) or viral inactivation with 2 mL of sample preservation solution (Mole BioScience, China) and RNA extraction, purification and concentration using an aliquot robot (Microlab^®^ STAR M, Hamilton Robotics, US) and reagents (MagMAX^®^ Viral/Pathogen Nucleic Acid Isolation kit, Thermofisher, US). RNA amplification was performed following the same procedure as direct RT-PCR.

#### Direct RT-qPCR intra- and inter-assay precision

A set of saliva specimens including one sample with high SARS-CoV-2 RNA load, one sample with low RNA load, one negative sample, and a negative control (water) were tested by triplicate in the same run to assess intra-assay precision. Three sets of saliva specimens including each of them one SARS-CoV-2 high positive sample, one low positive sample, one negative sample, and a negative control were tested in different runs in different days to evaluate inter-assay precision.

#### Effect of saliva storage conditions on direct RT-qPCR performance

SARS-CoV-2 RNA detection yield by direct RT-qPCR was determined for different conditions of saliva storage: at room temperature for a maximum period of 24 hours, refrigerated at 4°C for 24 hours, or frozen −80°C for longer than 24 hours.

### Diagnostic validation procedures

Diagnostic validation was conducted using samples collected from participants in the ongoing “Kids Corona Study of SARS-CoV-2 transmission at Football Club Barcelona Academy “La Masia”, run by SJDH. In brief, that study entailed self-collection of saliva by teen and young adult soccer, basketball, handball, futsal, and roller hockey players, as well as adult acompanying coaches, teachers, physiotherapists, and staff residing at or attending the Football Club Barcelona Academy “La Masia” (Barcelona, Spain). A team of SJDH research nurses supervised saliva self-collection by participants on site and collected paired NP swabs from them for comparative testing. Inclusion criteria in the diagnostic validation process were participant recruitment during August 2020 and follow up for at least 9 weeks. Collected saliva and NP samples were transferred to sterile Eppendorf tubes and NP VITM tubes respectively, labelled, and transported by the nurses in ambient temperature to SJDH’s Biobank for storage (saliva) or to SJDH’s Molecular Microbiology Department (NP samples) for standard RT-qPCR. Saliva was self-collected at baseline and on a weekly basis whereas NP samples were collected at baseline and every second week. Serum-based enzyme-linked immunoassays (ELISA) were also performed at baseline. All baseline saliva, NP, and serum samples were tested at study start and any saliva and NP samples paired with ELISA-positive specimens were excluded from the validation. In case of a positive RT-qPCR result in a NP sample, both the paired biobanked saliva sample collected at the same time point and the series of saliva samples obtained previously from the same participant were retrieved and retrospectively analysed by direct RT-qPCR using GeneFinder COVID-19 Plus RealAmp kit. Results by any RT-PCR method were interpreted as positive if at least two target genes of SARS-CoV-2 were detected and the amplification curves were adequate; and inconclusive if either only one gene was detected or amplification curves were unusual.

### Pilot screening procedures

#### Saliva self-collection coaching

Health workers, students, aid volunteers, and other professionals of the study setting were invited to participate in a pilot screening programme for SARS-CoV-2 based on the validated method. Instructions were disseminated to participants so that they could collect their own saliva in an unsupervised but safe manner. Participants were recommended to collect their own saliva in the first morning hours or after a fasting period of 2 hours to avoid food remains, according to recent evidence [17]. They were instructed to spit their saliva into tube collectors, transfer samples to sterile Eppendorf tubes with disposable Pasteur pipettes, close tubes with screw caps, decontaminate external surfaces of tubes with a hydroalcoholic solution, and identify them with heat resistant barcode labels before delivery to the SJDH Molecular Microbiology Department. All the information about the adequate pre-analytical procedure was gathered in an explanatory video and a brochure. This training material was made accessible on line to the participants through SJDH’s intranet web site.

#### Pilot screening programme

Eppendorf tubes received in the laboratory were not opened until the virus had been inactivated with heat, for safety reasons. A high throughput system was put into service for rapid screening workflow utilising an aliquot robot (Microlab^®^ STAR M, Hamilton Robotics, US) and a thermal cycler (QuantStudio 7^®^, Thermofisher, US). Up to 384 batched RNA extracts, positive, and negative controls were dispensed by the aliquot robot to the PCR plate of the thermal cycler for performance of direct RT-qPCR reaction with TaqPath COVID-19 RT-PCR kit reagents. This process workflow can be completed in less than 2 hours.

### Statistical analysis

SARS-CoV-2 detection yields in saliva by direct and standard RT-qPCR, measured in cycle threshold (Ct) values, were compared using the Student t-test or the Mann-Whitney U test. Mean Ct values were determined from the SARS-CoV-2 genetic loci targeted by the two commercial RT-PCR kits used. Differences between Ct values obtained for SARS-CoV-2 targeted genes in different replicates and runs were analysed to assess precision. Associations between Ct values of direct RT-qPCR in saliva samples exposed to different storage temperature conditions were evaluated using the Pearson coefficient of correlation. Diagnostic sensitivity, specificity, positive and negative predictive values were determined as reported elsewhere [18]. Statistical significance was set at a p-value of <0.05 and confidence intervals (CI) at 95% level. All statistical analyses were performed using Stata v.15 software (Stata Corp., TX, US).

## Results

### Analytical validation results

A higher mean SARS-CoV-2 Ct value was obtained in saliva using GeneFinder amplification reagents for direct RT-qPCR (28.2, SD 6.0) compared to standard RT-qPCR (24.9, SD 5.4, *p*=0.13). In contrast, mean Ct values yielded by direct RT-qPCR when utilising TaqPath reagents were lower for direct RT-qPCR (21.4, SD. 5.2) than for standard RT-qPCR (22.8, SD 6.4, *p*=0.76). A significant difference was observed in mean Ct values yielded by direct RT-qPCR depending on the commercial reagents used (TaqPath, 21.4; GeneFinder, 28.2, *p*<0.01). The difference in mean Ct values for standard RT-qPCR did not significantly vary by commercial kit (TaqPath, 22.8; GeneFinder, 24.9, *p*=0.34) (Table 1). Saliva-based direct RT-qPCR showed differences in Ct value in a range of −0.99 to 2.84 within a run of replicates (Table 2) and in a range of −5.57 to 4.28 between diferent runs (Table 3). Strong correlations were found between Ct values for samples stored at room temperature, in refrigerator for 24 hours or frozen at −80°C, either for direct RT-PCR using GeneFinder kit (Pearson coefficient of correlation r>0.99) or TaqPath kit (r>0.94) (Table 4).

**Table 1.** Cycle threshold values of direct RT-qPCR and standard RT-qPCR in saliva.

|  | GeneFinder COVID-19 Plus RealAmp kit |  |  |  | TaqPath COVID-19 RT-PCR kit |  |  |  |
| --- | --- | --- | --- | --- | --- | --- | --- | --- |
|  | SARS-CoV-2 gene | Direct RT-qPCR (A) | Standard RT-qPCR<sup>a</sup> (B) | Difference (A-B) | SARS-CoV-2 gene | Direct RT-qPCR (C) | Standard RT-qPCR<sup>b</sup> (D) | Difference (C-D) |
| Sample 1 | *IC (RnasaP)* | 29.30 | 26.63 | 2.67 | *IC (MS2)* | - | 32.57 | - |
|  | *E* | 20.25 | 15.09 | 5.16 | *S* | 11.62 | 11.99 | -0.37 |
|  | *N* | 18.22 | 15.23 | 2.99 | *N* | 13.36 | 11.52 | 1.84 |
|  | *R* | 20.35 | 15.58 | 4.77 | *ORF1ab (R)* | 11.23 | 11.10 | 0.13 |
| Sample 2 | *IC (RnasaP)* | 28.73 | 25.94 | 2.79 | *IC (MS2)* | 23.95 | 24.09 | -0.14 |
|  | *E* | 30.15 | 26.56 | 3.59 | *S* | 23.04 | 23.74 | -0.70 |
|  | *N* | 28.28 | 26.21 | 2.07 | *N* | 24.73 | 24.83 | -0.10 |
|  | *R* | 29.09 | 26.83 | 2.26 | *ORF1ab (R)* | 22.71 | 23.02 | -0.31 |
| Sample 3 | *IC (RnasaP)* | 27.97 | 25.35 | 2.62 | *IC (MS2)* | 28.44 | 24.01 | 4.43 |
|  | *E* | 27.20 | 24.27 | 2.93 | *S* | 22.46 | 22.05 | 0.41 |
|  | *N* | 26.15 | 24.25 | 1.90 | *N* | 23.67 | 23.53 | 0.14 |
|  | *R* | 26.60 | 24.73 | 1.87 | *ORF1ab (R)* | 23.75 | 22.24 | 1.51 |
| Sample 4 | *IC (RnasaP)* | 29.53 | 25.81 | 3.72 | *IC (MS2)* | 34.70 | 23.70 | 11.00 |
|  | *E* | 28.71 | 27.58 | 1.13 | *S* | 19.74 | 26.30 | -6.56 |
|  | *N* | 27.82 | 27.18 | 0.64 | *N* | 23.59 | 26.75 | -3.16 |
|  | *R* | 28.33 | 28.39 | -0.06 | *ORF1ab (R)* | 22.17 | 25.60 | -3.43 |
| Sample 5 | *IC (RnasaP)* | 26.87 | 24.76 | 2.11 | *IC (MS2)* | 27.67 | 23.71 | 3.96 |
|  | *E* | 41.24 | 31.32 | 9.92 | *S* | 24.54 | 29.97 | -5.43 |
|  | *N* | 34.26 | 29.18 | 5.08 | *N* | 28.15 | 30.01 | -1.86 |
|  | *R* | 36.05 | 31.00 | 5.05 | *ORF1ab (R)* | 26.21 | 29.28 | -3.07 |

<sup>a</sup> SARS-CoV-2 RNA inactivation, extraction and amplification by NucliSense easyMag reagents and platform
<sup>b</sup> SARS-CoV-2 RNA preservation by Mole Bioscience, RNA extraction by MagMAX Viral/Pathogen Nucleic Acid Isolation and Microlab STAR M platform and RNA amplification by TaqPath COVID-19 reagents and Thermofisher thermal cycler

**Table 2.** Cycle threshold values of direct RT-qPCR in saliva replicates.

| Direct RT-qPCR result<sup>a,b</sup> | SARS-CoV-2 gene | Replicate 1 (A) | Replicate 2 (B) | Replicate 3 (C) | Difference (A-B) | Difference (A-C) | Difference (B-C) |
| --- | --- | --- | --- | --- | --- | --- | --- |
| Negative | *IC (RnasaP)* | 23,40 | 23,35 | 23,40 | 0,05 | 0,00 | -0,05 |
|  | *E* | - | - | - | - | - | - |
|  | *N* | - | - | - | - | - | - |
|  | *R* | - | - | - | - | - | - |
| Low positive | *IC (RnasaP)* | 23,44 | 23,45 | 23,40 | -0,01 | 0,04 | 0,05 |
|  | *E* | 37,93 | 37,10 | 35,09 | 0,83 | 2,84 | 2,01 |
|  | *N* | 31,84 | 32,83 | 31,58 | -0,99 | 0,26 | 1,25 |
|  | *R* | 32,23 | 32,89 | 32,03 | -0,66 | 0,20 | 0,86 |
| High positive | *IC (RnasaP)* | 23,21 | 23,31 | 23,29 | -0,10 | -0,08 | 0,02 |
|  | *E* | 23,36 | 23,31 | 23,67 | 0,05 | -0,31 | -0,36 |
|  | *N* | 23,01 | 22,91 | 23,23 | 0,10 | -0,22 | -0,32 |
|  | *R* | 23,77 | 23,58 | 24,01 | 0,19 | -0,24 | -0,43 |

<sup>a</sup>Direct-qPCR was performed after heat treatment using GeneFinder COVID-19 Plus RealAmp reagents and Thermofisher thermal cyclers
<sup>b</sup>The three replicates yielded negative results for the negative controls

**Table 3.**
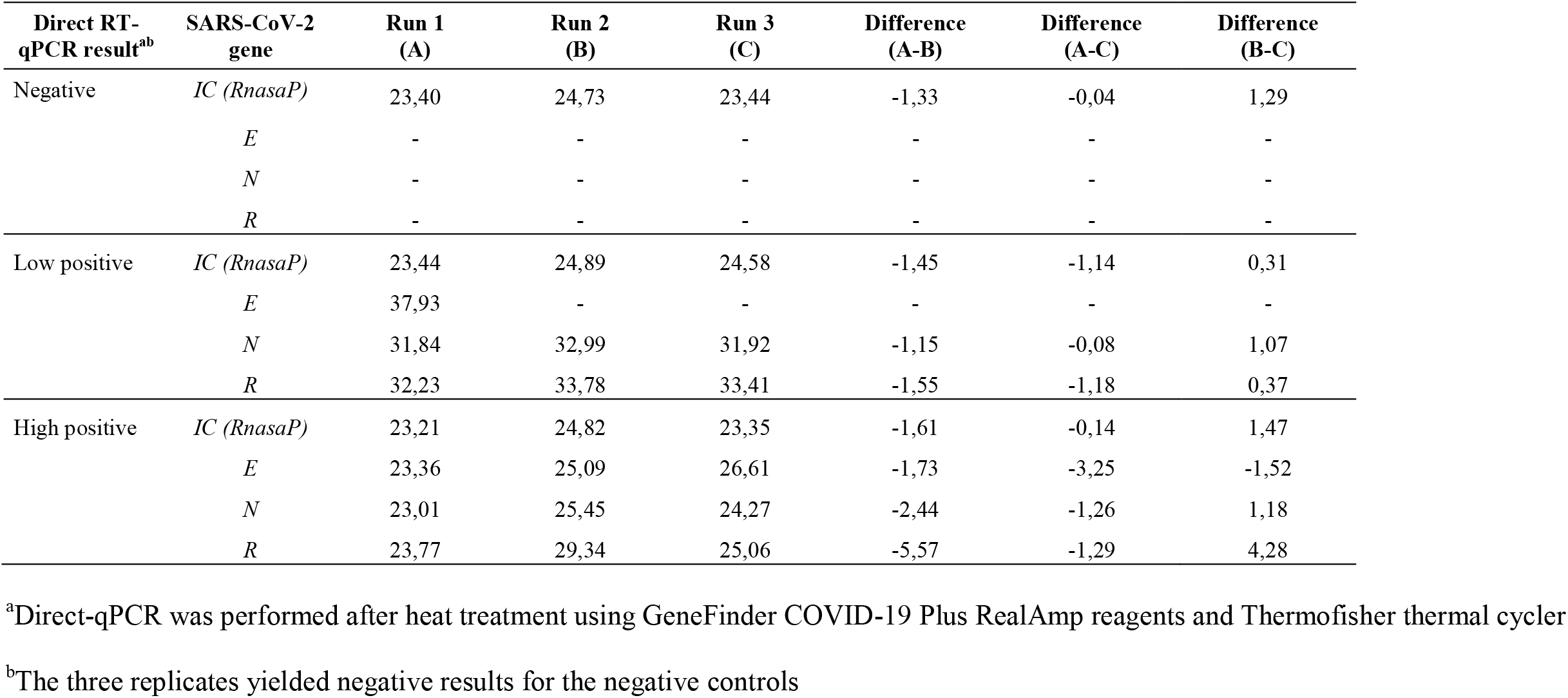
Cycle threshold values of direct RT-qPCR in saliva runs.

| Direct RT-qPCR result<sup>a,b</sup> | SARS-CoV-2 gene | Run 1 (A) | Run 2 (B) | Run 3 (C) | Difference (A-B) | Difference (A-C) | Difference (B-C) |
| --- | --- | --- | --- | --- | --- | --- | --- |
| Negative | *IC (RnasaP)* | 23,40 | 24,73 | 23,44 | -1,33 | -0,04 | 1,29 |
|  | *E* | - | - | - | - | - | - |
|  | *N* | - | - | - | - | - | - |
|  | *R* | - | - | - | - | - | - |
| Low positive | *IC (RnasaP)* | 23,44 | 24,89 | 24,58 | -1,45 | -1,14 | 0,31 |
|  | *E* | 37,93 | - | - | - | - | - |
|  | *N* | 31,84 | 32,99 | 31,92 | -1,15 | -0,08 | 1,07 |
|  | *R* | 32,23 | 33,78 | 33,41 | -1,55 | -1,18 | 0,37 |
| High positive | *IC (RnasaP)* | 23,21 | 24,82 | 23,35 | -1,61 | -0,14 | 1,47 |
|  | *E* | 23,36 | 25,09 | 26,61 | -1,73 | -3,25 | -1,52 |
|  | *N* | 23,01 | 25,45 | 24,27 | -2,44 | -1,26 | 1,18 |
|  | *R* | 23,77 | 29,34 | 25,06 | -5,57 | -1,29 | 4,28 |

<sup>a</sup>Direct-qPCR was performed after heat treatment using GeneFinder COVID-19 Plus RealAmp reagents and Thermofisher thermal cycler
<sup>b</sup>The three replicates yielded negative results for the negative controls

**Table 4.** Cycle threshold values of direct RT-qPCR in saliva according to storage conditions.

| GeneFinder COVID-19 Plus RealAmp kit |  |  |  |  |  |  | TaqPath Covid-19 COVID-19 RT-PCR kit |  |  |  |  |  |  |
| --- | --- | --- | --- | --- | --- | --- | --- | --- | --- | --- | --- | --- | --- |
| SARS-CoV-2 gene | Room temperature, 24 hours (A) | Refrigeration at 4°C, 24 hours (B) | Freezing at -80°C, >24 hours (C) | Difference (A-B) | Difference (A-C) | Difference (B-C) | SARS-CoV-2 gene | Room temperature, 24 hours (A) | Refrigeration at 4°C, 24 hours (B) | Freezing at -80°C, >24 hours (C) | Difference (A-B) | Difference (A-C) | Difference (B-C) |
| *IC (RnasaP)* | 29,70 | 31,65 | 31,68 | -1,95 | -1,98 | -0,03 | *IC (MS2)<sup>a</sup>* | - | - | - | - | - | - |
| *E* | 20,25 | 19,57 | 20,44 | 0,68 | -0,19 | -0,87 | *S* | 11,62 | 13,67 | 13,21 | -2,05 | -1,59 | 0,46 |
| *N* | 18,22 | 19,33 | 18,22 | -1,11 | 0,00 | 1,11 | *N* | 13,36 | 13,66 | 12,9 | -0,30 | 0,46 | 0,76 |
| *R* | 20,35 | 19,57 | 20,37 | 0,78 | -0,02 | -0,80 | *ORF1ab (R)* | 11,23 | 13,33 | 13,11 | -2,10 | -1,88 | 0,22 |
| *IC (RnasaP)* | 28,73 | 28,03 | 27,97 | 0,70 | 0,76 | 0,06 | *IC (MS2)<sup>a</sup>* | - | - | - | - | - | - |
| *E* | 30,15 | 30,50 | 30,56 | -0,35 | -0,41 | -0,06 | *S* | 23,04 | 24,33 | 24,6 | -1,29 | -1,56 | -0,27 |
| *N* | 28,28 | 28,54 | 28,51 | -0,26 | -0,23 | 0,03 | *N* | 24,73 | 25,47 | 25,51 | -0,74 | -0,78 | -0,04 |
| *R* | 29,09 | 29,31 | 29,33 | -0,22 | -0,24 | -0,02 | *ORF1ab (R)* | 22,71 | 25,3 | 25,08 | -2,59 | -2,37 | 0,22 |
| *IC (RnasaP)* | 27,97 | 27,77 | 27,81 | 0,20 | 0,16 | -0,04 | *IC (MS2)<sup>a</sup>* | - | - | - | - | - | - |
| *E* | 27,20 | 27,57 | 27,53 | -0,37 | -0,33 | 0,04 | *S* | 22,46 | 22,97 | 22,17 | -0,51 | 0,29 | 0,80 |
| *N* | 26,15 | 26,38 | 26,42 | -0,23 | -0,27 | -0,04 | *N* | 23,67 | 23,35 | 22,72 | 0,32 | 0,95 | 0,63 |
| *R* | 26,60 | 26,90 | 26,96 | -0,30 | -0,36 | -0,06 | *ORF1ab (R)* | 23,75 | 19,85 | 22,69 | 3,90 | 1,06 | -2,84 |

<sup>a</sup>Internal control of TaqPath COVID-19 RT-PCR kit was not tested because it is included directly in the RT-qPCR reagent mix

### Diagnostic validation results

A total of 183 out of 230 participants in the “Kids Corona Study of SARS-CoV-2 transmission at Barça” (185 children, 45 adults) met inclusion criteria. Ten participants were excluded from the validation process because they were positive for SARS-CoV-2 antibodies by ELISA at baseline. The remaining 173 participants yielded negative results in both paired saliva and NP samples at baseline and were followed up during 9 to 12 weeks. A positive RT-PCR in NP sample was detected in 23 participants in weeks 4 (n=1), 6 (n=1), 9 (n=4), 10 (n=7), 11 (n=2), and 12 (n=8). SARS-CoV-2 positivity was confirmed by direct RT-qPCR in 22 paired saliva samples and one was inconclusive. Of note, viral RNA was detected in the saliva samples of three participants one week earlier than being detected for the first time in NP specimens (Fig 1). An inconclusive result in NP specimen by standard RT-qPCR was obtained for seven participants. All serial paired (n=100) and non-paired preceeding saliva samples obtained from these subjects were found negative by direct RT-qPCR, except for one individual whose paired saliva yielded a positive result. Sensitivity and specificity values were 95.7% (95% CI, 79.0-99.2%) and 100.0% (95% CI, 98.6-100.0 %), respectively (Table 5). Positive predictive value was 100.0% (95% CI, 85.1-100.0%) and negative predictive value was 99.6% (95% CI, 98.0-99.9%).

**Table 5.** Positive, negative, and inconclusive results in paired saliva-nasopharyngeal samples.

|  |  | Saliva-based direct RT-qPCR |  |  | Total |
| --- | --- | --- | --- | --- | --- |
|  |  | Positive | Negative | Inconclusive |  |
| Standard RT-qPCR<br>in NP sample | Positive | 22 | 0 | 1 | 23 (7.6%) |
|  | Negative | 0 | 273 | 0 | 273 (90.1%) |
|  | Inconclusive | 1 | 6 | 0 | 7 (2.3%) |
|  | Total | 23 (7.6%) | 279 (92.1%) | 1 (0.3%) | 303 (100.0%) |

**Figure 1.**
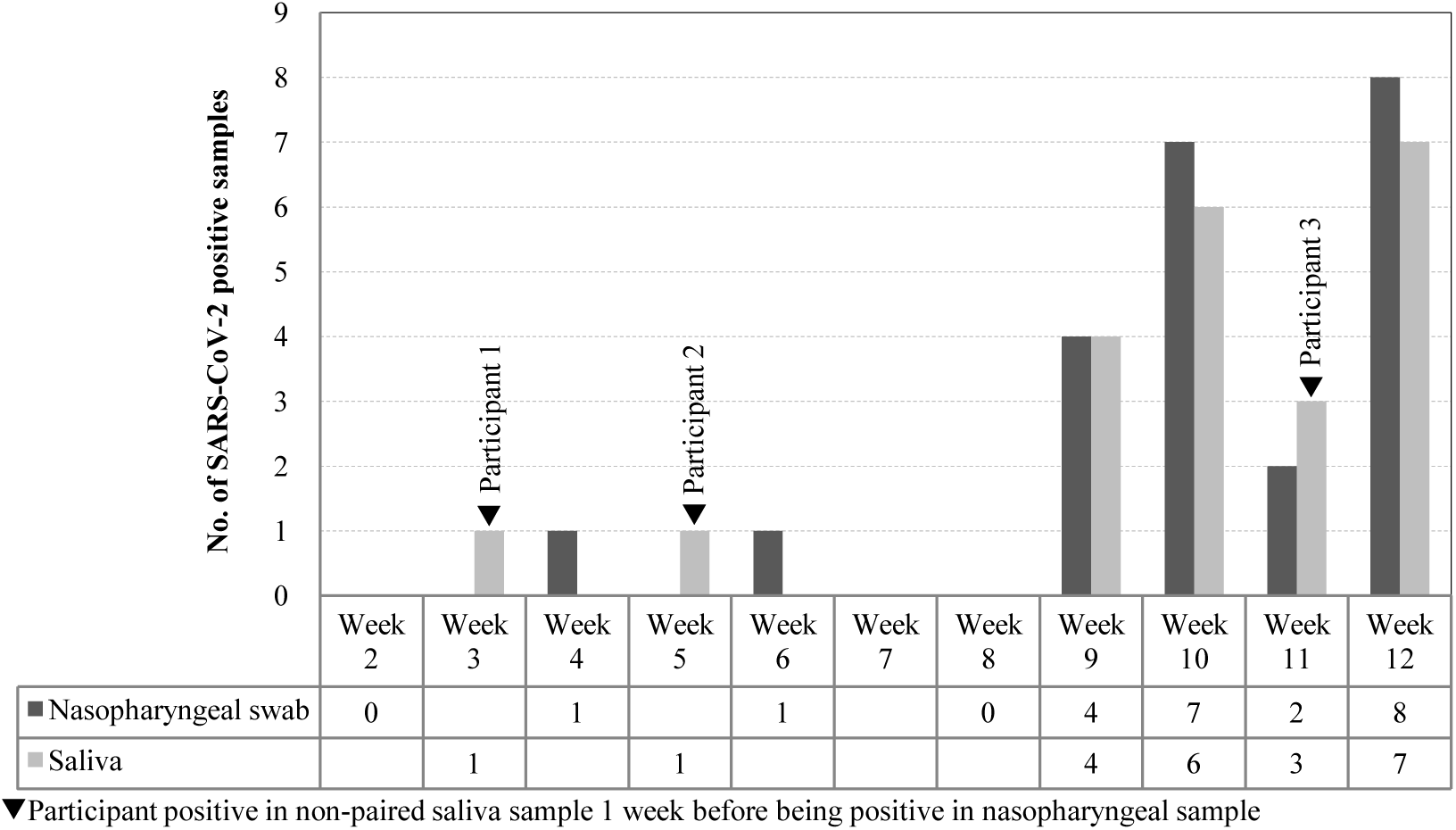
Time distribution of first positive SARS-CoV-2 result by sample type.

### Pilot screening programme

A total of 2,709 symptomless participants voluntarily engaged in the SARS-CoV-2 pilot screening programme in SJDH during 10 labour days of October 2020, including 2,076 (83.4%) out of 2,489 total health workers, 203 students, 23 aid volunteers, and other 407 professionals. Seventeen (0.6%) saliva samples provided by participants yielded invalid results by direct RT-qPCR. Among the remaining 2,692 saliva specimens, direct RT-qPCR was positive in 24 (0.9%) and inconclusive in 27 (1.0%). NP swabs were collected from participants with positive or inconclusive saliva results and tested by standard RT-PCR. All 24 (100.0%) participants with saliva-positive results were also found positive by standard RT-qPCR in NP swab. Four (14.8%) out of 27 participants with inconclusive saliva results were positive by RT-qPCR in NP swab and 23 were negative.

## Discussion

There is a lack of evidence on feasibility and usefulness of saliva-based RT-qPCR protocols for early SARS-CoV-2 infection. This study reports the results of validation and subsequent implementation of a direct RT-qPCR method based on end-user self-collection of raw saliva. Despite by-passing use of VITM and RNA extraction reagents, this method achieved high accuracy for screening asymptomatic individuals. Sensitivity (95.7%) and specificity values (100.0%) validated in a diverse cohort of teenagers and young and older adults without symptoms were comparable to those of standard RT-qPCR protocols that use NP samples for clinical diagnosis. Of note, the only saliva result discrepant from a positive result in NP sample was inconclusive. Thus direct RT-qPCR in saliva flagged the need of confirmatory testing for the individual with this inconclusive saliva result and fulfilled its screening purpose. Interestingly, we identified three subjects in the validation cohort that were positive in saliva one week before giving a positive result in NP sample. Since subjects were screened in saliva weekly and in nasopharynx every second week, this finding suggests that serial screening for SARS-CoV-2 should not consider frequencies longer than one week between successive tests to be effective.

When the method was implemented for pilot screening of SARS-CoV-2 in a reference hospital, all saliva-positive results (0.9%) agreed with positive results in paired NP samples. In addition, a few inconclusive results in saliva (1.0%) raised the need for confirmatory testing and uncovered a minor proportion of additional NP positive samples. Overall, these results indicate that the proposed method performs adequately in a real-life scenario for its intended use of screening. It is worthwhile to highlight that no significant usability issues occurred during the pre-analytical phase, as shown by the negligible proportion of invalid results obtained in saliva (0.6%). Moreover, pilot screening gained high participation among health workers in the study site, suggestive of their willingness to self-collect and dispense saliva samples according to a simple set of instructions. In operational terms, use of a high throughput system allowed fast analytical workflow for close surveillance and timely control of potential SARS-CoV-2 nosocomial infection in the setting. Overall, we speculate that method implementation may result in savings both in consumables (swabs, PPEs, VITM, RNA extraction reagents) and health workforce before RNA amplification step.

Research on SARS-CoV-2 RNA detection in pre-heated URT specimens other than saliva has been addressed by diverse groups, with a primary focus on diagnosis of symptomatic patients [19-21]. A pre-print study by Fernández-Pittol and colleagues specifically compared accuracy of direct RT-qPCR on saliva against standard RT-PCR on NP or oropharyngeal swabs of adult patients with SARS-CoV-2 symptoms [22]. While we observed 95.7% sensitivity and 100% specificity of saliva-based direct RT-qPCR in asymtomatic individuals, that group reported sensitivity and specificity values of 90.0% and 87.5%, respectively, in a cohort of adults who had experienced symptoms onset within the preceding 9 days. Being the protocols of heat treatment identical in both studies, our hypothesis is that after SARS-CoV-2 acquisition, viral load in saliva may progressively decrease across the pre-symptomatic stage, a declining trend that could continue after onset of symptoms, as already observed in symptomatic patients [23,24].

Few studies have analysed performance of saliva-based direct RT-qPCR on individuals without SARS-CoV-2 symptoms. In this regard, our results were consistent with those of a study by Wyllie and colleagues with 495 asymptomatic health workers tested by a RT-qPCR protocol that included use of VTM and previous RNA extraction [23]. This study found 13 saliva samples positive as well as all of their paired NP samples. In contrast, Williams et al., in their study of ambulatory patients attending a screening clinic, reported a lower saliva performance in 33 (84.6%) out of 39 self-collected saliva samples paired with 39 NP-positive samples [24]. They used a RT-qPCR protocol preceeded by RNA extraction from saliva diluted in Amiens medium and did not specify the proportion of asymptomatic infection in recruited outpatients. Similarly, a pre-print study by Kojima et al. determined SARS-CoV-2 RNA positivity rates of 79% in saliva and 85% in NP samples among a group of symptomatic and asymptomatic individuals (approximate ratio 1:1) who tested positive by at least one of various URT samples [25]. Their RT-qPCR workflow included saliva collection swabs, VTM use, and RNA extraction. Differences in pre-analytical steps prior to RT-qPCR and in proportions of asymptomatic individuals studied could explain the different performance of saliva-based RT-qPCR reported in our study and in William’s and Kojima’s.

A number of studies available in preprint have addressed development and clinical validation of saliva-based direct RT-PCR methods for SARS-CoV-2 screening, yet results of their implementation are unknown. Ranoa et al. developed a method that includes heat inactivation at 65°C during 30 minutes and use of sample stabilizing buffers (Tris-EDTA and Tris-Borate-EDTA) and additives (Tween 20) to enhance detection [26]. This group reported high sensitivity (88.9%) and specificity (98.9%) values in 9 paired positive and 91 paired negative saliva-NP samples. Similarly, Vogels et al. developed a protocol that consists of mixing saliva without preservative buffers with a proteinase K before performing heat inactivation at 95°C during 5 minutes and a dualplex RT-qPCR [27]. High positive (97.1%) as well as negative agreement (100.0%) was found in 37 paired positive and 91 paired negative saliva-NP samples.

Comparatively, our optimized method did not require addition of specific buffers to saliva for optimal performance while maintaining process workflow as safe and simple as possible.

The main strengths of this study were diagnostic validation of the proposed method in a diverse cohort of asymptomatic teenagers and young and older adults, as well as extensive method implementation for screening SARS-CoV-2 in a hospital environment. A limitation was that significant differences in Ct values were observed for direct RT-qPCR depending on the use of GeneFinder or TaqPath amplification reagents in the analytical validation process. To be noted, GeneFinder kit is designed for performance of 45 amplification cycles whereas TaqPath kit entails 40 cycles, and each of them sets different threshold values set for a positive result (GeneFinder, 40; TaqPath 37). Therefore, we were not able to provide insights into the significance of saliva viral load or Ct values obtained from these two commercial reagent kits.

In conclusion, this study showed that a novel direct RT-qPCT on self-collected raw saliva is a simple, safe, and accurate method for first-line screening of SARS-CoV-2. High throughput pilot implementation proved to be feasible, allowed fast analytical workflow, and gained high levels of voluntary participation in a sensitive hospital scenario. Self-collection of saliva by end-users had negligible effects on validity of results. Evidence generated by this study supports the potential scale up of self-collected, saliva-based direct RT-qPCR for enhanced community-wide screening of SARS-CoV-2.

## Data Availability

The data that support the findings of this study are available on reasonable request from the corresponding author. The data are not publicly available due to [restrictions e.g. their containing information that could compromise the privacy of research participants].

## Author summary

### Why dis study was done?

➢ Saliva is a promising non-invasive specimen type for screening, diagnosis, follow up, and infection control of SARS-CoV-2 in a safe and convenient manner.
➢ Diverse studies have addressed performance of RT-qPCR in saliva for clinical diagnosis of symptomatic patients. However, there is scarce evidence about saliva-based RT-qPCR protocols aimed to screen asymptomatic subjects.

### What did the researchers do and find?

➢ We validated and implemented an optimised screening method that combines use of self-collected raw saliva samples, single-step heat-treated virus inactivation and RNA extraction, and direct RT-qPCR.
➢ Saliva-based RT-qPCR showed high sensitivity (95.7%) and specificity (100.0%) to identify asymptomatic individuals in a validation cohort of 173 teenagers and young and older adults, compared to standard RT-qPCR in nasopharyngeal sample.
➢ A high throughput pilot screening of 2,709 staff in a sensitive reference hospital scenario gained high levels of participation (83.4% among health workers). The pilot proved feasibility of unsupervised self-collection of saliva by participants (only 0.6% of invalid results) and potential for rapid analytical workflow (up to 384 batched samples can be processed in <2 hours).
➢ All participants screened as positive in saliva (n=24) were also positive by standard RT-qPCR in nasopharyngeal sample. Four out of 27 participants with inconclusive saliva results were flagged and also confirmed as positive.

### What do these findings mean?

➢ The present study serves as a demonstration that direct RT-qPCT on self-collected raw saliva is a simple, rapid, and accurate method that can be scaled up for enhanced community-wide screening of SARS-CoV-2.

## Acknowledgements

Wxe are indebted to SJDH’s Biobank (Cristina Jou, Anna Codina), Occupational Health and Prevention Department (Pilar Subirats and Olga Nadal), Clinical Laboratory Department (Cristina Esteva, Assumpta Fasanella, Manuel Monsonis, and Ana Valls) and “Kids Corona Study Group” (Mariona F de Sevilla, Claudia Fortuny, Maria Mele, Maria Rios, Laia Alsina, Elisenda Bonet, Aleix Garcia-Miguel, Iris Uribesalgo, Cristina Jou, Anna Codina, Maite Miranda, Felipe Perez-Soler and Marta Cubells) of SJDH. We also thank Dr. Jordi Vila from Hospital Clinic Barcelona and Dr. Dirk Egging and Dr.Adam Meijer from National Institute for Public Health and the Environment from The Neetherlands for sharing their experience on RT-PCR in saliva.

## Contributors

PB and CMA designed the study and wrote the paper. APA and JS performed experiments and collected data. ST, GR collected data. PB, CL, VF, EG, IJ, QB, and CMA analysed and interpreted data. JC, JGG, IJ and GR managed participant recruitment. CMA supervised the study. All authors discussed the results and critically reviewed, discussed and accepted the final version of the manuscript. The corresponding author attests that all listed authors meet authorship criteria and that no others meeting the criteria have been omitted.

## Ethical approval

The study was approved by the Ethics Commitee of SJDH prior to the beginning of activities (ref. PIC-240-20). Use of samples collected from participants in the “Kids Corona Study of SARS-CoV-2 transmission at Football Club Barcelona Academy “La Masia” for the present and future studies was covered in the informed consent process and approval of that study (ref. PIC-200-20).

